# Questionable practices in data and code sharing policy in high-profile medical journal and research

**DOI:** 10.1101/2024.08.29.24312818

**Authors:** Wei Li, Xuerong Liu, Qianyu Zhang, Liping Shi, Jing-Xuan Zhang, Xiaolin Zhang, Jia Luan, Yue Li, Ting Xu, Rong Zhang, Xiaodi Han, Jingyu Lei, Xueqian Wang, Yaozhi Wang, Hai Lan, Xiaohan Chen, Yi Wu, Yan Wu, Lei Xia, Haiping Liao, Chang Shen, Yang Yu, Xinyu Xu, Chao Deng, Pei Liu, Zhengzhi Feng, Chun-Ji Huang, Zhiyi Chen

## Abstract

**Background:** The spurious and unavailable data/code sharing actions are crashing open medical sciences. In this study, we aimed to illustrate how high-profile medical journals are practically carried out their sharing policies and what questionable practices regarding data/code sharing are conducted by authors.

**Methods:** In this study, we appraised the policy on data/code availability of high-profile medical journals ranked at Q1 according to Clarivate Journal Citation Report (JCR 2021). Furthermore, we recruited post-publications published by four leading medical journals (i.e., The BMJ, JAMA, NEJM and The Lancet) from the issuing of data/code availability policy to December 2022 for the questionable practices in data/code sharing. The appraisal of papers was conducted by the Data/code Availability Statement Practice Evaluation Tool (DANCE), developed by systematically integrating mainstreaming open data/code guidelines.

**Findings:** We found that less than one-tenth journals (9.1%) mandated authors to share data/code, with an available statement. Among these journals, 70.6% (61.2%) did not consider censoring (restricting) spurious/invalid data/code sharing in publications. Furthermore, though journal impact factor could predict policy stringency on “offering availability statements” (*p* < .001), it failed to predict ones in “sharing data/code” (*p* = .73). For publications, even in leading medical journals (i.e., The BMJ, JAMA, NEJM and The Lancet), only 0.5% of the papers (16/3,191) fully complied with their public sharing statements for reaching reproducibility. Lack of availability statement, declining data/code sharing without reasons, and invalid repositories were leading questionable practices conducted by authors.

**Interpretation:** We clarified specific questionable actions of implementing and practicing the sharing policy both in journal and papers, which should be addressed not only by the supportive publication ecosystem but also by crediting authors for taking responsibility and maintaining scientific integrity in data/code sharing.

**Funding:** No funding.

## Introduction

The importance of data and code sharing is evident, as it has been well-acknowledged to benefit the best interests of science and public health good, particularly in promoting open, transparent, reproducible and trustworthy sciences^1, 2^. Despite such promising and ambitious goals, the actions toward these grand promises are seriously challenged^3, 4^. Though declaring to embrace and welcome open data/code sharing initiatives/ethics, the prevalence of issuing data sharing policies in (even leading) medical journals (i.e.,< 10%) has not yet achieved open science goals, with no prominent changes resulting from the implementation of institutional or stakeholder-driven data sharing statements in the last decade^5^. Moreover, in these journals with policies requiring or even mandating sharing, 98.0% (99.5%) of papers do not actually share available research data (code), showing a notorious gap between declared policies and actual publication practices^6^. However, for these challenges, we know very little about how these journal policies are actually carried out, and what questionable practices are exactly conducted by authors in these publications.

Journals are the first gatekeeper to prevent spurious or unavailable data/code sharing practices. Many scientific communities such as The Transparency and Openness Promotion (TOP)^7^ and the Recommendation on Open Science from United Nations Educational, Scientific and Cultural Organization (EOP-UNESCO)^8^ offer structural guidelines to formulate open data/code policies at the journal side, clearly indicating step-by-step censorship to data/code sharing integrity with certified standards (e.g., FAIR data stewardship)^9^. However, neither supportive policy changes nor increased priority to data sharing practices have occurred after proclaiming these guidelines. Beyond such community-based recommended guidelines, the International Committee of Medical Journal Editors (ICMJE) has ever imposed to report data sharing statement/plan to clinical trial data^10^, but nothing has changed in the prevalence of data sharing^11^. Despite the consensus on poor prevalence, it has still been underexplored regarding how journals practically carry out their sharing polices. For example, *did journals merely require a statement to report data sharing methods, but not require actual data sharing? or did high-rank journals guarantee stringent policy on data/code sharing?*

For the policy-to-practice gap, though spurious and actually unavailable data/code sharing statements are incredibly pervasive^6, 12^, we can do nothing to address this notorious deterioration until we understand what (intentionally or inadvertently) questionable practices are conducted by authors^13^. On the one hand, clarifying these specific questionable research actions may contribute to guiding journals in tailoring sharing policies, with add-on clauses, to self-correct policy loopholes for actually valid and available sharing^14^. Moreover, such findings may additionally benefit editorial or peer review in censoring data/code sharing integrity, indicating notable risky points in their data/code sharing statements^15^. On the other hand, for authors, identifying reasons of incurring failure on actually available data/code sharing in their publication practices could further educate authors on how to adjust sharing strategies when the journal policy is not yet supportive/helpful enough. Supporting this argument, the poor data/code sharing implementation has been partly attributed to the low practicability in these guidelines/policies per se^16^.

Here, to answer the first question, we utilized the data/code availability policy (DAP) matrix (**Table 1**) to appraise degree of policy on data/code availability in 931 high-profile medical journals ranked at Q1 according to Clarivate Journal Citation Report (JCR 2021). To answer the second question, for the four leading medical journals (i.e., British Medical Journal, BMJ; Journal of the American Medical Association, JAMA; New England Journal of Medicine, NEJM; Lancet), we evaluated 3,191 papers published from the issuing of data/code availability policy to December 2022 for the questionable practices in data/code sharing/availability. This meta-research appraisal was conducted by the Data/code Availability Statement Practice Evaluation Tool (DANCE), which was developed by systematically integrating mainstreaming open data/code guidelines **(eTables 2-6)**.

**Table 1:** The Data/code Availability Policy (DAP) matrix. The degree of data/code availability policy of journal primarily depends on the requirements regarding data sharing policy and data availability statement policy. Note: both “optional” and “encouraged” in data/code sharing policy and availability policy refer to that the willingness of sharing data or providing availability statement depends on the authors themselves, but differing in the strength of the suggestion, with “encouraged” being more strongly worded than “optional”.

|  | Data/code Availability Statement Policy |  |  |
| --- | --- | --- | --- |
|  | No policy | Optional/Encouraged | Required |
| **Data/code Sharing Policy** |  |  |  |
| No policy | No policy | Encouraged | Required |
| Optional/Encouraged | Encouraged | Encouraged | Required |
| Required | Encouraged | Encouraged | Mandated |

## Methods

### Study design

The current study employed meta-research to appraise the implementation of high-profile medical journal policies on data/code sharing and availability statements. Furthermore, the study also aimed to assess questionable data/code sharing risks and to identify specific questionable practices impeding data/code sharing in papers published in leading medical journals.

### Search strategy and eligibility criteria

For journal policies, we began by selecting all medical journals ranked at Quartile 1 (Q1) in the category of Clinical Medicine from Clarivate Journal Citation Report (JCR) 2021 on 1st June 2023. Journals publishing original research were included in the present study, while review journals and book series were excluded. We finally included 931 journals for the appraisal of policies.

For data/code availability practices in papers, we first included all papers published in the leading journals including the British Medical Journal (The BMJ), Journal of the American Medical Association (JAMA), New England Journal of Medicine (NEJM) and The Lancet, restricting the time from issuing sharing policy (2018 for BMJ and JAMA; 2019 for NEJM; 2021 for The Lancet) to 2022. All of the leading journals required authors to provide an availability statement to disclose whether the data/code would be shared for specific paper types. All specific article types which were required to provide such statement by journals were included, such as original articles and brief reports, while case report, review, abstract and qualitative research were excluded. Finally, 3,191 papers were included in the current analysis.

Two reviewers (L.W. and L.X.R.) independently checked the eligibility for the included journals and papers by screening the journals’ aim, scope and guidelines for authors, as well as all papers’ types, titles, abstracts and main texts. Any disagreements were resolved by a senior reviewer (C.Z.Y.).

### Data collection and processing

For eligible journals, the journal characteristics, including Uniform Resource Locator (URL), 2021 impact factor, publisher, subspecialty, publication frequency, International Standard Serial Number (ISSN), and electronic ISSN, were firstly extracted from Clarivate Journal Citation Report (JCR 2021) by two reviewers (L.W. and L.X.R.). For journals belonging to multiple subspecialties, we counted them in each subspecialty, in the subgroup analysis of subspecialties. Then, two reviewers (Z.Q.Y. and W.Y.Z.) systematically extracted the descriptions regarding the data/code sharing and availability statement recorded in Guidelines for Authors manually for appraising the implementation of journal policies.

For eligible papers, two reviewers (H.X.D. and L.J.Y.) manually extracted bibliographic characteristics (journal, title, published year, article type, DOI, study design) and data/code availability statement for assessing questionable data/code sharing risks and to identify specific questionable practices impeding data/code sharing as guided by the DANCE tool (see below “**Data Analysis**”).

### Data Analysis

In the analysis of journals’ policies, we built on a data/code availability policy (DAP) matrix (**Table 1**). The degree of each journal was classified into four ranks based on the policy description regarding data/code sharing and availability statement. “No policy” denotes the complete absence of any directives or guidelines regarding data/code sharing and availability statements. “Encouraged” denotes that the decision to share data/code or provide an availability statement is left to the discretion of the authors. “Required” denotes that the journal requires authors to include an availability statement specifying whether data/code will be shared. “Mandated” denotes that the journal obliges authors to both share data/code and provide an availability statement detailing the methods through which the shared data/code can be accessed by others. The degree of each journal depends on the description of policies. We further conducted subgroup analysis of journals’ policies, including subspecialties, publishers and journal impact factors. Three reviewers (X.X.Y., D.C. and L.P.) independently rated the policies, with disagreement solved by the fourth author (C.Z.Y.).

In the analysis of papers’ practices, the Data/code Availability Statement Practice Evaluation Tool (DANCE) was developed by systematically to assessing integrating and structuring mainstream open data/code guidelines, which was developed to assess questionable data/code sharing risks and to identify specific questionable practices impeding data/code sharing, from four domains based on the process of reproducing the results of papers **(eTables 2-6)**. The first domain (statement integrity) refers to the comprehensiveness of the statement, ensuring it encompasses all key elements as stipulated by the journal’s policy. The second domain (actual accessibility) refers to the practical implementation of the availability statement, confirming that the data/code are indeed accessible as originally claimed. The third domain (user usability) refers to the retrieved data/code are organized in a manner that facilitates user-friendly verification. The fourth domain (method practicability) refers to whether the original results can be fully reproduced utilizing the accessed data/code. In each domain, reviewers are required to answer several signaling questions with “yes”, “probably yes”, “no”, “probably no” or “unclear/not applicable”. Each domain was rated as high risk of questionable data/code sharing practices if 1 or more items were answered with no/probably no. Only applicable articles could be rated in each domain. For example, when a paper declares not to share its data/code, it is not applicable to Domains 2,3, and 4, and would not be rated in these domains. Nine authors (L.W., L.X.R, Z.Q.Y., L.J.Y., H.X.D., W.Y.Z., W.X.Q., S.C. and Y.Y.) independently rated the policies, with disagreement solved by the tenth author (C.Z.Y.).

### Statistical analysis

We utilized descriptive statistical analyses in Microsoft Excel 2021, presenting these data as frequencies and rates. Furthermore, in the subgroup analysis of journal impact factor, we used Spearman correlation analysis conducted by SPSS (IBM, Inc., version 29.0.1.0).

## Results

### Journal policies on data/code sharing

Of 931 journals, 42.2% (393/931) required a statement disclosing data/code availability, 28.7% (267/931) did not require it (i.e., encouraged/optional), and 29.1% (271/931) did not include any clauses (**Fig. 1a**). For data/code sharing policy, more than half of the journals (601/931, 64.6%) did not require authors to share data/code (i.e., encouraged/optional), 22.6% (210/931) did not include any clauses, and only 12.9% (120/931) required data/code sharing (**Fig. 1 b**). In summary, in these high-profile medical journals claiming embrace of open data initiatives/ethics, less than one-tenth of the journals (85/931, 9.1%) mandated authors to share research data/code along with a clear availability statement (**Fig. 1 c**). To make matters worse, even among the 85 journals with mandatory data availability policies, 60 journals (70.6%) had not yet disclosed any implemented measures in their policies to ensure that data/code shared in publications were genuine and valid, and 52 journals (61.2%) had not considered making sharing of spurious or invalid data/code a disqualifying condition for publication.

**Fig. 1:**
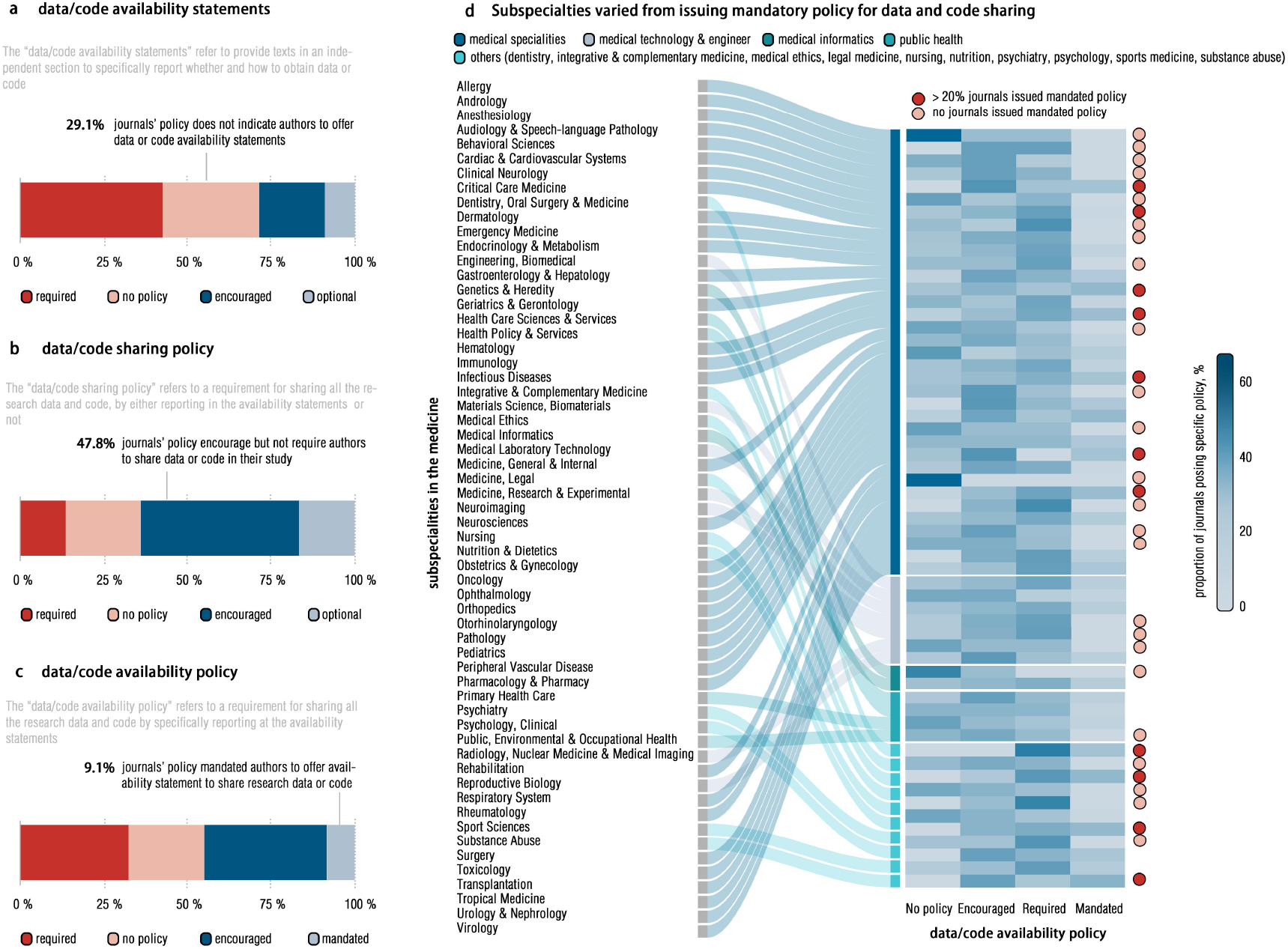
The proportions of policy degrees for all Q1 journals regarding data/code availability. (a) shows the degree of the requirement for data/code availability statement in publications. (b) shows the degree of the requirement for data/code sharing in publications. (c) shows the overall degree concerning both availability statement and data/code sharing. (d) illustrated the distribution of overall degree across subspecialties (n=59) in medical science. For journals belonging to multiple subspecialties, we counted them in each subspecialty.

In subgroup analyses, by categorizing into subspecialties and publishers, we further examined the above proportions of journals’ policies across 59 medical subspecialties and 13 publishers (containing at least 10 journals). Regarding the subspecialties, subspecialties with over 20% of journals implementing mandatory data/code sharing and statement policies accounted for 18.6% (11/59) (**Fig. 1 d**). As for the publishers (**Fig. 2 a**), the proportion of such mandatory policies varied largely from 0% to 50%. Finally, we conducted Spearman correlation analysis to investigate whether the 2021 Journal Impact Factors (JIFs) could predict the policy compliance. Interestingly, though the journal ranks quantified by 2021 JIFs could predict policy stringency on “offering data availability statement” (ρ=0.20, 95% CI: 0.13-0.26, p<0.001), it failed in predicting the policies on “sharing actual data/code” (ρ=0.01, 95% CI: -0.06-0.08, p=0.737; **Fig. 2 b**), which possibly implied the risk of the practice of formalities for formalities’ sake in promoting research transparency.

**Fig. 2:**
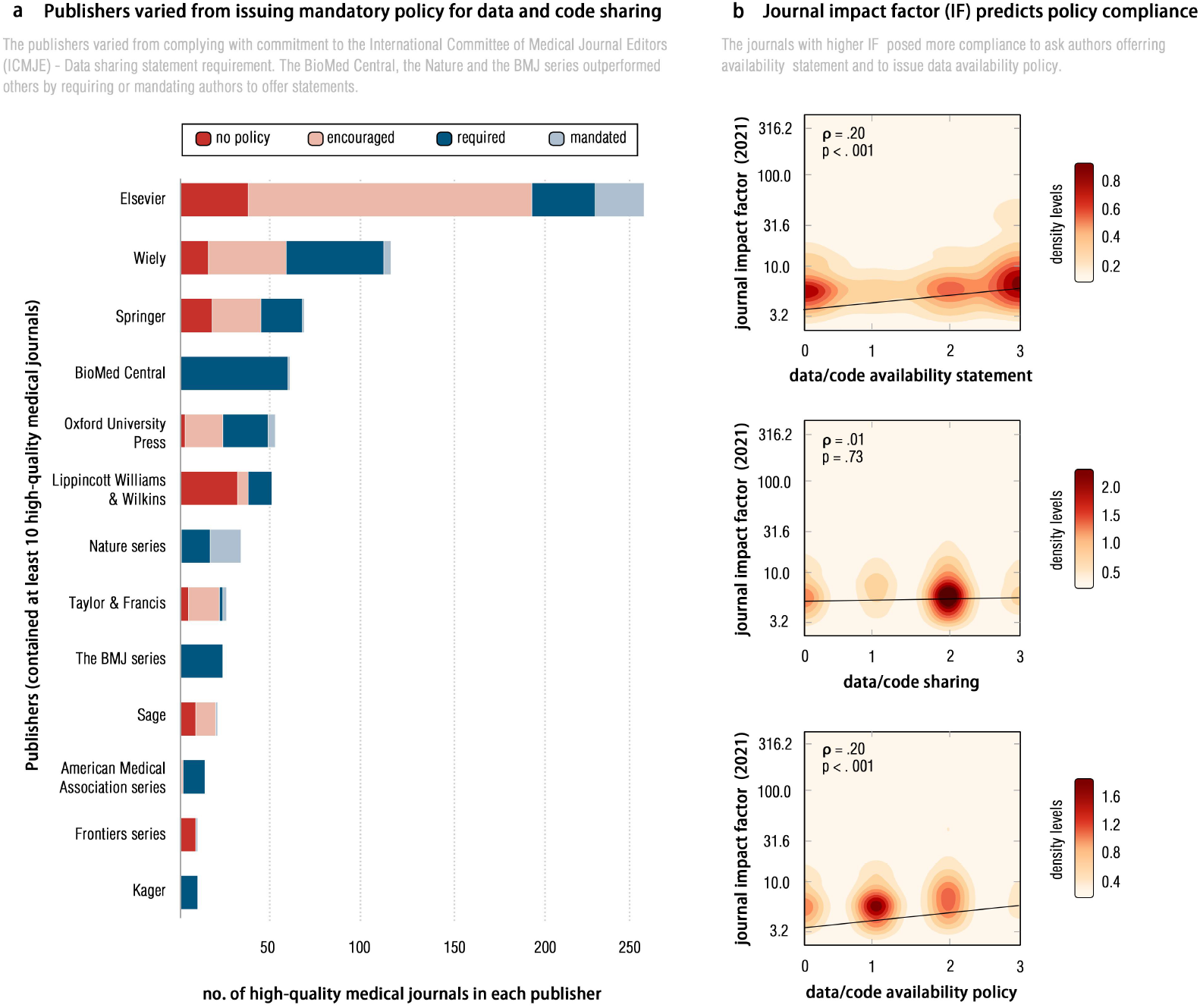
The subgroup analyses in publishers and journal impact factors. (a) shows the distribution of overall degree across publishers. We only included publishers containing at least 10 high-profile medical journals. (b) shows the correlation between the policy of data/code availability statement, data/code sharing or overall degree (data/code availability) and journal impact factor (2021) according to the Clarivate Journal Citation Report (JCR) 2021, estimated by Spearman Correlation Analysis.

### The risk of conducting questionable data/code sharing

When examining data/code sharing statements for individual 3,191 papers, 92.3% (1,555/1,685) of those with private availability and 95.8% (413/431) with public availability were rated as high risk for questionable sharing practices. The remaining 33.7% (1,075/3,191) papers lacked the statements. For each specific appraisal domain in the DANCE **(Table 2)**, 93.3% were rated as high risk for statement integrity, 46.1% for actual accessibility, 89.5% for user usability, and 76.5% for method practicality. Consequently, only 0.5% (16/3,191) could be empirically reproduced by following their sharing statements.

**Table 2:** The risk of questionable data/code sharing practices for papers using Data/code Availability Statement Practice Evaluation Tool (DANCE) (N=3,191). Only applicable articles could be rated in each domain. For example, when a paper declares not to share its data/code, it is not applicable to Domains 2,3, and 4, and would not be rated in these domains. Each domain was rated as high risk of questionable data/code sharing practices if 1 or more items were answered with no/probably no. a. low risk of questionable data/code sharing practices. b. high risk of questionable data/code sharing practices. Articles that were not applicable were excluded from the risk proportion calculations. c. fully reproduced papers.

|  | Yes/probably yes | No/probably no | Unclear/Not Applicable |
| --- | --- | --- | --- |
| **Domain 1: Statement Integrity** | *213 (6.7%)<sup>a</sup>* | *2,978 (93.3%)<sup>b</sup>* | 0 |
| 1.1 Was this paper provided Data Availability Statement or/and Code (Scripts) Availability Statement or/and Materials Availability Statement. | 2,124 (66.6%) | 1,067 (33.4%) | 0 |
| 1.2 Was this paper explained why data, code (scripts) or materials sharing were under restrictions when it claimed “upon reasonable request” or “no additional data available”? | 952 (29.8%) | 567 (17.8%) | 1,672 (52.4%) |
| 1.3 Were these data, code (scripts) or materials deposited in a reliable repository, with specific hyperlinks or entrances? | 179 (5.6%) | 170 (5.3%) | 2,842 (89.1%) |
| 1.4 Was this repository has unique and persistent identifiers (e.g., DOI)? | 181 (5.7%) | 1 (0.0%) | 3,009 (94.3%) |
| 1.5 Was this Data Availability Statement or/and Code (Scripts) Availability Statement or/and Materials Availability Statement discoverable? | 2,124 (66.6%) | 0 | 1,067 (33.4%) |
| 1.6 Were the data, code (scripts) or materials sharing integral? | 248 (7.8%) | 1,857 (58.2%) | 1,086(34.0%) |
| **Domain 2 Actual Accessibility** | *104 (53.9%)<sup>a</sup>* | *89 (46.1%)<sup>b</sup>* | 2,998 |
| 2.1 Were these hyperlinks or entrances valid actually? | 176 (5.5%) | 17 (0.5%) | 2,998 (94.0%) |
| 2.2 Were the permissions or accesses actually given for all the users? | 112 (3.5%) | 64 (2.0%) | 3,015 (94.5%) |
| 2.3 Were these accessing data, code (scripts) or materials actually conformed to statement? | 109 (3.4%) | 8 (0.3%) | 3,074 (96.3%) |
| 2.4 Were these accessing data, code (scripts) or materials actually obtained? | 115 (3.6%) | 0 | 3,076 (96.4%) |
| 2.5 Were these obtained files workable? | 114 (3.6%) | 1 (0.0%) | 3,077 (96.4%) |
| **Domain 3: User Usability** | *12 (10.5%)<sup>a</sup>* | *102 (89.5%)<sup>b</sup>* | 3,077 |
| 3.1 Were these accessed data/code/materials indicated for how to use? | 42 (1.3%) | 72 (2.3%) | 3,077 (96.4%) |
| 3.2 Were these accessed data/code/materials organized user-friendly? | 108 (3.4%) | 6 (0.2%) | 3,077 (96.4%) |
| 3.3 Were the software or statistical toolkit prepared to use these data/code/materials? | 62 (1.9%) | 52 (1.6%) | 3,077 (96.4%) |
| 3.4 Were these data/code/materials understandable for all the users (e.g., junior, trained and senior researchers)? | 13 (0.4%) | 101 (3.2%) | 3,077 (96.4%) |
| **Domain 4: Method Practicability** | *16 (23.5%)<sup>a</sup>* | *52 (76.5%)<sup>b</sup>* | 3,123 |
| 4.1 Were the programming or technical environments generalizable for users? | 67 (2.1%) | 1 (0.0%) | 3,123 (97.9%) |
| 4.2 Were these codes/scripts actually workable? | 12 (0.4%) | 12 (0.4%) | 3,167 (99.2%) |
| 4.3 Were the necessary comments or information were provided within code or scripts? | 54 (1.7%) | 4 (0.1%) | 3,133 (98.2%) |
| 4.4 Were the dependent software/packages/toolkit provided? | 59 (1.8%) | 8 (0.3%) | 3,124 (97.9%) |
| 4.5 Were these code/scripts editable? | 11 (0.3%) | 14 (0.4%) | 3,166 (99.2%) |
| Reproducibility | 16 (0.5%)<sup>c</sup> |  |  |

### Specific questionable practices impeding data/code sharing

Given the pervasive risk of questionable data/code sharing identified above, We synthesized questionable practices from “high-risk” papers and found that one-third of the papers (1,075 of 3,191, 33.7%) did not include any descriptions about data/code availability, despite the clear requirements of availability statements by journals **(Fig. 3)**. Though providing such statements, 23.3% (745/3,191) refused to share data/code, in which 58.4% (435/745) did not provide any reasons. Even worse, 13.5% (431/3,191) declared to share data/code publicly, but 39.0% (168/431) were not reachable to the sharing data/code. The remaining 29.5% (940/3,191) papers declared to share data/code upon request, with neither clarification to permission conditions nor explanations to the data/code preservation.

**Fig. 3:**
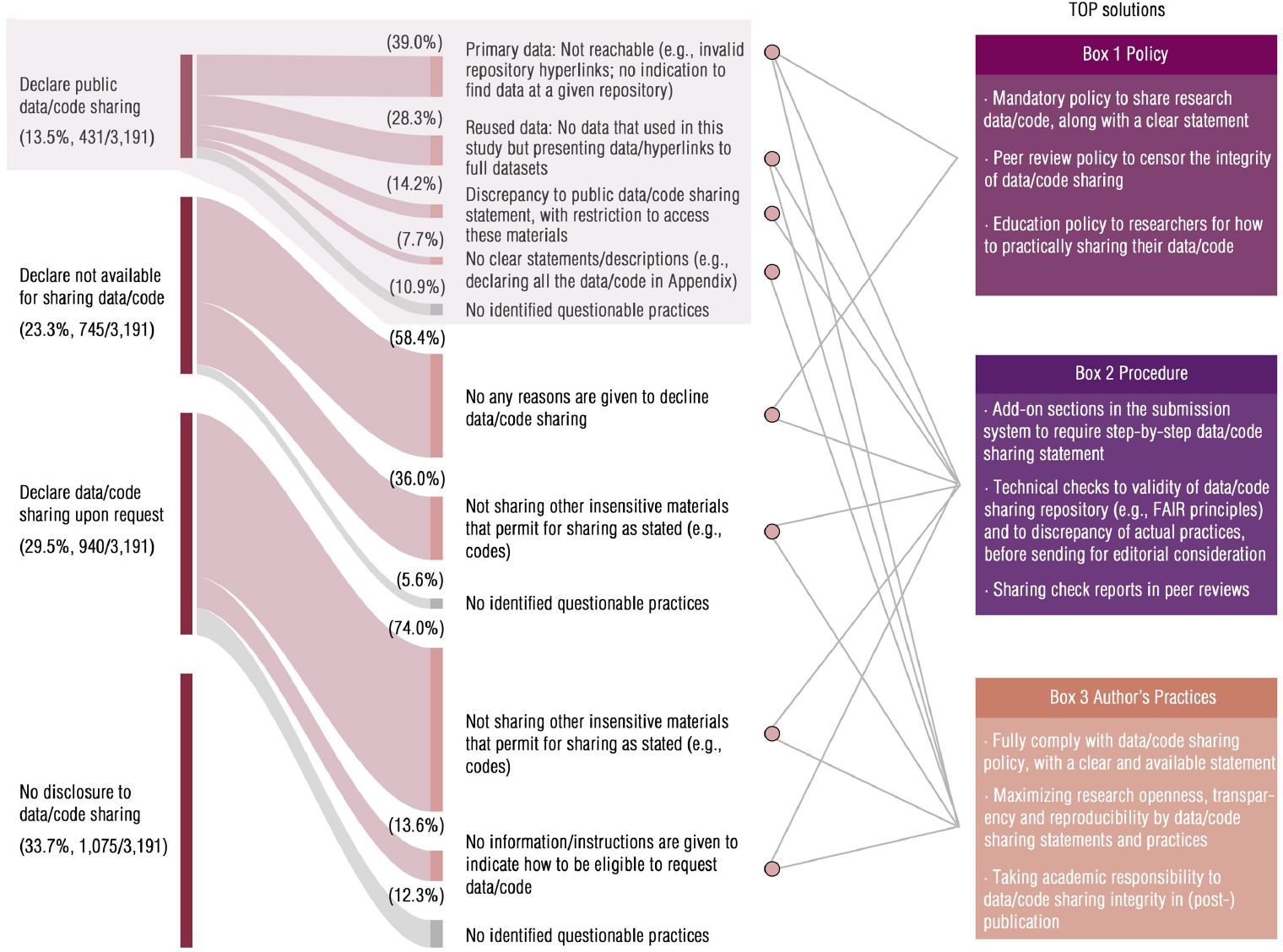
The questionable data/code availability practices in publications and solutions according to the Transparency and Openness Promotion (TOP) Guideline. The practices were structured based on the declared data/code availability and actual data/code availability within each type of declared availability. The TOP solutions were divided into journal policy, submission procedure, and author’s practices as outlined in the TOP Guideline.

## Discussion

Data and code sharing policy certainly fueled open, transparent and better science, but not as much as we expected. In this meta-research study, we found the journal policies and grandiloquent promises did not guarantee scientific integrity on open science practices. Even in the high-profile medical journals, less than one tenth of them declared a mandate for authors to share data/code, along with a clear availability statement. Moreover, these journals neither stipulated censorship of these statements (70.6%) nor enforced valid data/code sharing as an obligatory condition of publication (61.2%). Even worse, such questionable practices in the journal policy on data/code sharing seemed to be pervasive, as we observed no significant associations between journal ranks (i.e., journal impact factor) and policy stringency. Compared to the motivation for declaring a data/code sharing policy, actual efforts to mandate or impose this practice on authors have been less prioritized. Beyond journal policy flaws, we found that authors (intentionally or inadvertently) conducted questionable practices on impeding valid data/code sharing, in leading medical journals, particularly in lacking sharing statements, declining data/code sharing without reasonable explanations, and invalidating data/code repositories. Despite the acknowledged low actual data/code sharing quality and low policy compliance^6, 12^, we exactly clarified what practices are conducted to hamper readers actually accessing their data/codes, which were suggested to be addressable at journal sectors or at authors per se.

For journal policy, though scientific communities and publishers vociferously promoted open science by supporting “data/code sharing policy”, our findings suggested that, most high-profile medical journals have merely taken “politically-correct” actions, with a formalistic (but not actual) requirements on policy practices. As previously reported, the editors and leading medical publishers (i.e., The Lancet) did not value or prioritize the quality of data/code sharing, as they have no editorial responsibilities for research practices but possess duties on publication ethics^17, 18^. Thus, such “meretricious” policies may readily become an “empty promise” to readers, for being posturing to endorse open sciences only^4^. Furthermore, though leading medical journals (e.g., The BMJ) indeed declared to take responsibility for data/code sharing^19^, they contributed not much to improve its validity, with less than 10% of these papers fully sharing research data/code. Therefore, it is evident that taking editorial responsibilities to implement mandatory policies alone is not enough to address questionable data/code sharing problems^20^. Instead, a supportive publication ecosystem including peer review, scientific integrity education, institutional funds, and technical censorship to data/code sharing practices is more imperative^21-23^.

For the overarching stakeholder - authors, despite their ostensible enthusiasm for open science, one disappointing truth is that many failed to responsibly share their data/code enough for reaching transparency and reproducibility. To our knowledge, beyond well-documented studies on the motivations impeding authors from sharing data/code (e.g., technical errors, high time costs, less credits/benefits, or competitive disadvantage)^24, 25^, this is the first study to clarify how authors may questionably practice data/code sharing policy in pursuing publication. One primary questionable conduct is not offering data/code sharing statements or restricting access upon request without reasonable considerations. This suggests that, rather than putting all trust on proactive actions from authors, compliance with data/code sharing policy should be prescribed as authors’ responsibility, which is a reflection of scientific integrity and even publication ethics per se^13, 26^. By doing so, the data/code sharing policy no longer earns open science credits for journals but directly benefits authors’ research integrity^27^. Furthermore, credit or incentive to such actions is the key^28^. As indicated by the TOP guideline, the “open data” badges, scientific integrity credits or other awards should be given to acknowledge that authors are taking the responsibility^29^. Another main questionable practice is to (intentionally or inadvertently) invalidate data/code sharing repositories while declaring public access. Technically speaking, this flaw could be readily addressed by technical scrutiny. For instance, an end-to-end text-mining model has been applied to automatically detect data sharing integrity, such as scrutinizing whether research data have been clearly claimed, fully accessed, actually downloaded and practically operated^30^. At authors’ side, crediting their responsibility and scientific integrity on data/code sharing, along with stringent technical checks, may be a practical solution.

Here, several limitations should be acknowledged. First, we narrowed the scope of probing data/code sharing policies to these high-profile medical journals, which alluded to the hypothesis that these were representative of stricter and high-quality policies in the medically science than others. Despite this premise has been substantially supported in previous evidences, a landscape of policy and its practices in all the medical journals (> 7000) could strengthen the reliability and generalizability of these findings. Second, we focused on the practical quality of public data/code sharing, and did not send data requests to validate practices in the statements declaring “upon request”. Third, given the lack of standardized and structural tool to systematically appraise practicability of data/code sharing, we tailored the DANCE by systematically integrating and structuring these mainstream open data/code guidelines. These findings could be further validated once a reliable checklist/tool was prepared.

In conclusion, journals’ policies to support transparent, open and better science are unprecedentedly ambitious and undoubtedly conducive to benefiting the medical community, but the actual effectiveness remains very suboptimal. Despite issuing data/code sharing policy, journals/publishers may merely focus on “declaring” rather “sharing” research data/code in the publication practices, without censorship and restriction to spurious/substandard “data availability” statements. Even in leading journals, under data sharing requirements in the policies, the questionable data/code sharing practices impeding data access in the papers are substantially pervasive. Authors may intentionally (or inadvertently) conduct multifarious questionable practices to repudiate data/code sharing. Such potential misconducts in practicing data/code sharing should have been addressed not only by establishing a supportive publication ecosystem but also by crediting authors for taking responsibility and maintaining scientific integrity in data/code sharing.

## Acknowledgments

we do thank Daniel G Hamilton for supports on data curation.

## Author Contributions

Zhiyi Chen, Xuerong Liu and Wei Li: Conceptualization, Methodology and Writing - Original Draft; Chun-Ji Huang, Jia Luan, Yue Li, and ZhengzhiFeng: Conceptualization; Liping Shi, Jing-Xuan Zhang, Ting Xu, Rong Zhang and Xiaolin Zhang: Writing - Review & Editing; Wei Li, Xuerong Liu, Qianyu Zhang, Xiaodi Han, Jingyu Lei, Xueqian Wang, Yaozhi Wang, Hai Lan, Xiaohan Chen, Yi Wu, Yan Wu, Lei Xia, Haiping Liao, Chang Shen, Yang Yu, Xinyu Xu, Chao Deng and Pei Liu: Data Curation & Validation; Zhiyi Chen: Project Administration and Funding Acquisition. L.W. and L.X.R. shared first authorship.

## Competing Interest Statement

All the authors disclosed no conflicts of interests.

## Data availability

The datasets of journal policy and article practice and the code required to reproduce all of the findings of this meta-research are available on the Open Science Framework (OSF) (https://osf.io/5m3zc/).

## Notes

### Competing Interest Statement

The authors have declared no competing interest.

### Funding Statement

This study did not receive any funding

