## Supplemental Table 1 for "Questionable practices in data and code sharing policy in high-profile medical journal and research"

**CONTENT**

**Supplementary Methods…………………………………………..……………………………………3**

1. **The data/code availability policy matrix………..........…………………………………………..3**
2. **Practices of Pursuing Reproducibility from “Data Availability Statements”……………....…3**
3. **Reference……………………………………………….……….………………………...…….….9**

|  | **Data/code Availability Statement Policy** | | |
| --- | --- | --- | --- |
|  | No policy | Optional/Encouraged | Required |
| **Data/code Sharing Policy** |  |  |  |
| No policy | No policy | Encouraged | Required |
| Optional/Encouraged | Encouraged | Encouraged | Required |
| Required | Encouraged | Encouraged | Mandated |

**The data/code availability policy matrix**

**eTable 1:** The Data/code Availability Policy (DAP) matrix. The degree of data/code availability policy of journal primarily depends on the requirements regarding data sharing policy and data availability statement policy. Note: both “optional” and “encouraged” in data/code sharing policy and availability policy refer to that the willingness of sharing data or providing availability statement depends on the authors themselves, but differing in the strength of the suggestion, with “encouraged” being more strongly worded than “optional”.

**Practices of Pursuing Reproducibility from “Data Availability Statements” in the Medical Science: A Meta-Research Evaluation**

1. Circuit for conducting a meta-research evaluation for the risk of low method reproducibility

| **Step** | **Task** | **Example** |
| --- | --- | --- |
| 1 | Specify your review topic (s) or/and targeting journal (s) | Neuroimaging-based diagnostic studies that published in Top 10% journals of Neuroimaging category (Clarivate JCR) |
| 2 | Specify your inclusion and exclusion criterion to obtain targeting paper (s) | Peer-reviewed English-writing papers; fMRI, MEG or fNIRs imaging techniques; Original studies |
| 3 | Assess risk of low reproducibility for each domain | Inadequate data availability information; invalid resources; unclear descriptions for how to use these resources |
| 4 | Overall evaluation of risk of low reproducibility and Data Availability Statement (DAS) practices | More than one answering “no” for any signaling question |
| 5 | Assess DAS practices for actual reproducibility | Independently reproducing all the analysis to replicate these findings basing on DAS |

**eTable 2:** The step for conducting a meta-research evaluation for the risk of low method reproducibility

2. Data Availability Statement Practice Evaluation Tool (DANCE)

**Domain 1: Statement Integrity**

| **Background**  Statement integrity could be flawed to incur risk of low reproducibility when the main contexts of this statement is incomplete or contradictory to journal policies or part criterion in the Guidelines for Transparency and Openness Promotion (TOP, v1.0.1). | |
| --- | --- |
| **1.1 Was this paper provided Data Availability Statement or/and Code (Scripts) Availability Statement or/and Materials Availability Statement.** | |
| Yes/Probably Yes | If this paper provided Data Availability Statement or/and Code Availability or/and Materials Availability at any sections or this paper implicitly indicated how to access them in the main texts of this paper. |
| No/Probably No | If this paper did not mentioned any information for how to obtain the data, code (scripts) or materials across the whole main texts. |
| Unclear | If no adequate information to make this judgement. |
| **1.2 Was this paper explained why data, code (scripts) or materials sharing were under restrictions when it claimed “upon reasonable request” or “no additional data available”?** | |
| Yes/Probably Yes | If this paper provided specific reasons to why restrict data, code (scripts) or materials sharing or/and provided how to access them under specific conditions; or if this paper had been provided specific information for data/code/materials accesses (see *1.1*). |
| No/Probably No | If this paper ONLY claimed “upon reasonable request” corresponding on any author (s) or “no additional data available”, without any specific explanations or conditions. |
| Unclear | If no adequate information to make this judgement. |
| **1.3 Were these data, code (scripts) or materials deposited in a reliable repository, with specific hyperlinks or entrances?** | |
| Yes/Probably Yes | If they were deposited in a trusted repository (e.g., Open Science Framework, Figshare) or institutional digital depository, with specific hyperlinks or entrances to (re-) visit them. |
| No/Probably No | If they were provided at personal store or website with temporary hyperlinks, entrances, or accesses. |
| Unclear | If no adequate information to make this judgement. |
| **1.4 Was this repository has unique and persistent identifiers (e.g., DOI)?** | |
| Yes/Probably Yes | If this repository have unique and persistent identifiers or labels. |
| No/Probably No | If this repository was not unique for this paper or had temporary identifiers or labels. |
| Unclear | If no adequate information to make this judgement. |
| **1.5 Was this Data Availability Statement or/and Code (Scripts) Availability Statement or/and Materials Availability Statement discoverable?** | |
| Yes/Probably Yes | If this statement was provided in an independent or special section; or if this statement was easily found in the main texts of this paper. |
| No/Probably No | If this statement was hidden in the supplementary information or elsewhere; of if this statement was hardly found in the main texts of this paper. |
| Unclear | If no adequate information to make this judgement. |
| **1.6 Were the data, code (scripts) or materials sharing integral?** | |
| Yes/Probably Yes | If this statement provided information for all the data (e.g., raw data, preprocessed data, formal analytic data and statistical data), all the code (scripts) and all the materials. |
| No/Probably No | If this statement provided information for a portion of data/code/materials; or if this statement provided complete data/code/materials for replicating partial results. |
| Unclear | If no adequate information to make this judgement. |

**eTable 3:** The evaluation items of Statement Integrity

**Domain 2: Actual Accessibility**

| **Background**  Flawed validity in the statement can be found when the actual practices in following this statement were not workable as original statement claimed. | |
| --- | --- |
| **2.1 Were these hyperlinks or entrances valid actually?** | |
| Yes/Probably Yes | If these hyperlinks or entrances linking with data/code/materials can be actually opened or accessed as DAS claimed; or if these hyperlinks or entrances were redirected to obtain them actually. |
| No/Probably No | If these hyperlinks or entrances linking with data/code/materials were no longer valid in any situations (e.g., 404 error, failure to load data). |
| Unclear | If no adequate information to make this judgement. |
| **2.2 Were the permissions or accesses actually given for all the users?** | |
| Yes/Probably Yes | If users could actually access all the data/codes/materials in this repository, without any restrictions, additional verification or permission requests; of if users need to freely register the personal account to access them, without any additional restrictions. |
| No/Probably No | If users must require a permission or access to obtain them; or if users could obtain partial data/codes/materials, with restrictions partial ones. |
| Unclear | If no adequate information to make this judgement. |
| **2.3 Were these accessing data, code (scripts) or materials actually conformed to statement?** | |
| Yes/Probably Yes | If these accessing data, code (scripts) or materials were fully consistent to what had claimed in the statement. |
| No/Probably No | If these accessing data, code (scripts) or materials were inconsistent to the statement; or if a portion of them were consistent with the statement only. |
| Unclear | If no adequate information to make this judgement. |
| **2.4 Were these accessing data, code (scripts) or materials actually obtained?** | |
| Yes/Probably Yes | If these accessing data, code (scripts) or materials were downloaded, transformed, obtained with reasonable ways. |
| No/Probably No | If these accessing data, code (scripts) or materials were obtained by technical constraints or unreasonable methods (e.g., ultra-low download speed, < 5kb/s; rare or specific downloader). |
| Unclear | If no adequate information to make this judgement. |
| **2.5 Were these obtained files workable?** | |
| Yes/Probably Yes | If these downloads were actually readable, workable and available. |
| No/Probably No | If these downloads were unworkable, unusable or unavailable. |
| Unclear | If no adequate information to make this judgement. |

**eTable 4:** The evaluation items of Actual Accessibility

**Domain 3: User Usability**

| **Background**  Risk of low reproducibility could occur when these accessed data/code/materials were hard to be used for verification. | |
| --- | --- |
| **3.1 Were these accessed data/code/materials indicated for how to use?** | |
| Yes/Probably Yes | If “Readme”, “Manual” or other guideline documents were provided to clearly indicate how to use them. |
| No/Probably No | If no any documents were provided to indicate what and how these files represented and used. |
| Unclear | If no adequate information to make this judgement. |
| **3.2 Were these accessed data/code/materials organized user-friendly?** | |
| Yes/Probably Yes | If they were deposited by a systematic/hierarchical framework (e.g., Raw Data, Preprocessed Data ...) or categorized manners (e.g., Result 1, Results 2 ...) |
| No/Probably No | If they were deposited by arbitrary orders; or if they were hard to be understood how to find specific components in their current forms. |
| Unclear | If no adequate information to make this judgement. |
| **3.3 Were the software or statistical toolkit prepared to use these data/code/materials?** | |
| Yes/Probably Yes | If software or specific/self-made/purpose-built toolkit were provided to use these data/code/materials; or if how to publicly obtain these software or toolkit when they are common or are under commercial conditions. |
| No/Probably No | If there were no sources to obtain software or toolkit; of if no information were provided to what software or toolkit should be applied for using them. |
| Unclear | If no adequate information to make this judgement. |
| **3.4 Were these data/code/materials understandable for all the users (e.g., junior, trained and senior researchers)?** | |
| Yes/Probably Yes | If they were introduced clearly for how to “quick start” or use “expert mode”; or if relevant documents were provided for step-by-step reproduction. |
| No/Probably No | If there were massive jargon, unclear abbreviation, or default parameters; of if no unclear/incomplete information were provided for how to reproduce these results by basic step-by-step manner. |
| Unclear | If no adequate information to make this judgement. |

**eTable 5:** The evaluation items of User Usability

**Domain 4: Method Practicability**

| **Background**  Risk of low reproducibility could occur when these accessed data/code/materials cannot verified from low method practicability, such as unreadable scripts, over-special programming environment and poor understandability. | |
| --- | --- |
| **4.1 Were the programming or technical environments generalizable for users?** | |
| Yes/Probably Yes | If the programming (e.g., R, Matlab, SPSS, Python) or technical environments (e.g., macOS, Linux) were general or common for broad users; of if users were instructed for how to set these specialized programming or technical environments |
| No/Probably No | If the programming or technical environments were too specialized/rare to be used for reproduction. |
| Unclear | If no adequate information to make this judgement. |
| **4.2 Were these codes/scripts actually workable?** | |
| Yes/Probably Yes | If they were successfully used by applying corresponding software or statistical toolkit. |
| No/Probably No | If they were not workable; of if the errors that were not addressable occur when using them; or if a portion of codes/scripts were workable only. |
| Unclear | If no adequate information to make this judgement. |
| **4.3 Were the necessary comments or information were provided within code or scripts?** | |
| Yes/Probably Yes | If necessary comments or information were provided to indicate how to use them and indicate what parameters were specialized within the code/scripts. |
| No/Probably No | If no any comments or inadequate information to indicate how to use them. |
| Unclear | If no adequate information to make this judgement. |
| **4.4 Were the dependent software/packages/toolkit provided?** | |
| Yes/Probably Yes | If necessary dependent software/package/toolkit were provided when using these code/scripts for reproduction. |
| No/Probably No | If a portion of dependent software/package/toolkit were provided only; or if dependent software/package/toolkit were indicated to be found elsewhere. |
| Unclear | If no adequate information to make this judgement. |
| **4.5 Were these code/scripts editable?** | |
| Yes/Probably Yes | If these code/scripts were editable for implementing both reproduction and replication. |
| No/Probably No | If editions for these code/scripts were limited partly or fully; of if these code/scripts could only editable to reproduce these specific parameters or the data. |
| Unclear | If no adequate information to make this judgement. |

**eTable 6:** The evaluation items of Method Practicability
